# Exploring the Roles of Male Partner in Transmission and Prevention of Cervical Cancer in Central Kenya: A Qualitative Approach

**DOI:** 10.1101/2025.05.09.25326715

**Authors:** J. H Mwangi, N. G Mtshali, P. N. Mbeje

**Affiliations:** School of Nursing and Public Health, University of KwaZulu-Natal, South Africa

**Author notes:** Corresponding Author: J.H Mwangi.

**Keywords:** Male partners involvement, Cervical cancer, transmission, prevention, Central Kenya

## Abstract

**Background:** Cervical cancer, a significant public health concern globally, arises from persistent infection with high-risk types of human papillomavirus (HPV). While the it primarily affects women, men play a crucial role in the transmission of HPV and also influence the processes of prevention and control of the disease.

**Aim:** The study aimed at exploring couples, health care workers and policy makers perceptions regarding the roles of male partner in the transmission, prevention, and control of cervical cancer.

**Setting:** The study was carried out in three public county hospitals located in Central Kenya.

**Methods:** The study used qualitative research design for a comprehensive exploration of the research topic. A total number of 73 participants who included 20 couples,20 Nurses two clinical officers two gynecologists, six Community health workers and three county directors of health were involved in the study. Thematic analysis was employed to identify recurring themes and patterns.

**Results:** Based on the participants perceptions, Male partners had limited knowledge about cervical cancer and were primarily influenced by myths and misconceptions. Their roles in transmission were in their behavior towards HPV spread while prevention and control included providing financial and logistical support, offering moral support, addressing HPV transmission and vaccination, navigating traditional and cultural practices, and contributing to health education and healthcare provision.

**Conclusion:** There is lack of awareness about cervical cancer. Financial constraints, cultural and social attributes and health care system influences males in supporting their partners in prevention and control of cervical cancer.

**Contribution:** Identifying the potential barriers and male partner role in encouraging his partner to go for cervical cancer screening.

## Background

While cervical cancer primarily affects women, men are not passive onlookers. Their actions and attitudes significantly influence the effectiveness of prevention efforts (Adegboyega et al., 2019). The study explored the perception of couples, health care workers and policy makers regarding male partner involvement in transmission and prevention of cervical cancer. The study also explored couples attitudes towards male partners support during cervical cancer prevention and control including their children HPV vaccination as sometimes there is HPV vaccination hesitancy due to lack of knowledge(Grandahl and Nevéus, 2021). Men can be asymptomatic carriers of HPV, meaning they may not experience any symptoms despite harboring the virus. This asymptomatic nature makes men unknowingly contribute to the spread of HPV through sexual contact. HPV can reside in the skin and mucosal tissues of the penis, urethra, and anus, enabling transmission during sexual activity (Castellsagué et al., 2003).Social factors significantly influence men’s role in HPV transmission. Cultural norms, societal expectations, and access to information about HPV and sexual health can shape men’s understanding of their role in transmission (Bishwajit et al., 2017). In some societies, men may have multiple sexual partners like those in polygamous relationships or the ones having concubines, increasing their risk of contracting and spreading HPV. Furthermore, limited access to comprehensive sexual education and healthcare services can hinder men’s awareness of HPV and its potential consequences (Lin et al., 2022).

Exploring strategies to augment knowledge about HPV and cancer screening and fostering trust in the healthcare system among male spouses or partners is essential, particularly with the aim of promoting cervical cancer awareness among immigrants in the USA (Read et al., 2020).Kenya cancer policy 2019-2030 have addressed many areas in regard to cancer prevention and management (MOH, 2020),however nothing is mentioned on partners support nor male input in prevention and controlling cervical cancer Studies demonstrate a favorable connection between men’s proactive engagement in reproductive healthcare and enhanced outcomes for maternal and child health. Increased male partner knowledge about cervical cancer will lead to increased uptake of cervical cancer screening and prevention services. (Bishwajit et al., 2017). To bridge this disparity, there is an urgent requirement for enduring community health education and promotion strategies that inspire and foster male involvement across various facets of reproductive healthcare, encompassing cervical cancer screening and treatment(Bola-Oyebamiji et al., 2024)

The study aimed at exploring the couples and health care workers perceptions and attitudes towards male partners contributions in transmission and prevention of cervical cancer. This can offer insight on what can be done to overcome challenges that may influence men participation in their partners journey of cervical prevention and control. Couples perceptions may indicate their knowledge (or lack of it) about cervical cancer prevention and control. Men’s sexual behaviors may directly contribute towards HPV transmission. Factors such as the number of sexual partners, condom use, and the age of sexual debut can influence their risk of contracting and spreading HPV (Cooper et al., 2018). Research indicates that men with multiple sexual partners are at a higher risk of acquiring and transmitting HPV. While condom use is not a guaranteed method of prevention, it can reduce the risk of HPV transmission (Pierce Campbell et al., 2013).

Men have a substantial influence on women’s decisions regarding reproductive health issues including cervical cancer prevention and control (Binka et al., 2019)(Sharma et al., 2018). Their involvement can positively impact women’s psychological, moral, and financial well-being, but it can also negatively affect them through stigmatization, isolation, or restrictions on healthcare access. Traditionally, sexual and reproductive health initiatives have focused primarily on women. However, recognizing the importance of male involvement in these discussions is crucial for promoting acceptance and care uptake (Adewumi et al., 2019). Research suggests that both women and men desire shared decision-making in reproductive matters, and a partner’s influence can significantly affect a woman’s adherence to reproductive health practices (Adewumi et al., 2019; Aborigo et al., 2018).

In many African cultures, men continue to hold a dominant role within family structures. This societal framework, emphasizing male leadership, can be leveraged to enhance women’s participation in cervical cancer screening programs (Adegboyega et al., 2019). To effectively promote cervical cancer awareness among immigrant populations in the USA, it was crucial to focus on strategies that increase male spouses or partners’ knowledge about HPV and cancer screening, as well as foster their trust in the healthcare system (Read et al., 2020). While the Kenya cancer policy for 2019-2030 addresses various aspects of cancer prevention and management (MOH, 2020), it notably lacks any mention of partner support or male involvement in preventing and controlling cervical cancer.

While the introduction of the HPV vaccine in certain regions of Kenya may have raised community awareness of cervical cancer, inadequate levels of knowledge and understanding could still lead to low screening rates among eligible women. Although various obstacles at the community, patient, and healthcare provider levels may impede screening, a comprehensive examination of these factors is currently lacking. These perceived obstacles include access (physical and financial, partner approval, stigma, embarrassment during screening, infertility due to the procedure, worry of residual effects of test results, lack of knowledge, and religious or cultural beliefs. (Buchanan Lunsford et al., 2017).

Spousal support in healthcare matters has a positive influence on health promotion and disease prevention, as highlighted by Adegboyega et al. (2019). By identifying the roles, patterns, and factors associated with male partner involvement in the transmission of cervical cancer and the uptake of screening and treatment services, we can improve the implementation of programs at the facility, county, and national levels in Kenya

In order to achieve herd immunity against HPV,the need to implement vaccination programmes that include all genders has been discussed yet few countries have included male children in their vaccination programmes (Milano et al., 2023).HPV is the most common sexually transmitted infection worldwide. There is a high detection rate in sexually active youth but the risk in males persist for many years. Various test have used for HPV detection and associated pathologies yet none has been approved for males (Vives et al., 2020).

### Research methods and design

A qualitative research approach (Cresswell, 2013)with Interpretivism and Constructivism ideologies was adopted to conduct the study. The interpretivism and constructivism paradigm advocate the notion that individuals are intentional and inventive in their actions, actively shaping their social environment. This approach recognizes the dynamic and evolving nature of society, acknowledging the possibility of multiple interpretations of an event influenced by individuals’ historical or social perspectives (Siddiqui, 2019, Ugwu et al., 2021)

The relevance of this approach to the study lies in the historical focus of reproductive health programs primarily on women as clients, overlooking the fact that women are not entirely independent of men. Women often have male figures in their lives, such as husbands, fathers, male relatives, and other significant others. Studies, such as the one conducted by Adegboyega et al. (2019), have shown that the support of spouses plays a crucial role in influencing the utilization of reproductive health services, particularly those pertaining to cervical cancer prevention and control programs It is imperative to get opinions of both genders regarding their perception on cervical cancer prevention and control. Male partners or men in general plays an important role in transmission, prevention and control of cervical cancer. The researchers contend that involving males as partners in the execution and delivery of cervical cancer screening and treatment services will contribute to an increased acceptance and utilization of these services.

A qualitative exploratory, descriptive and contextual research design (Hunter et al., 2019) was employed to explore the roles of male partners as perceived by couples, health care workers and policy makers. The design enabled the participants to share their opinions and experiences regarding male partner involvement in prevention and control of cervical cancer by giving them a platform to express their opinions and experiences.

### Study setting

This study was conducted at three county referral hospitals with MCH clinics in Central Kenya: Murang’a, Nyeri, and Kirinyaga.The selection of these three study areas was purposeful, driven by their high prevalence and incidence of cervical cancer, as indicated by hospital-based registries, in comparison to the national figures (Mwangi et al., 2017, MOH, 2015) As per the 2019 population census, the three counties have a population of 2,426,215, with 1,200,239 males and 1,225883 females, with a population density of 415/km^2^.

### Study participants and sampling strategy

The population of the study were 73 participants. These included 20 couples of reproductive age (18-50 years), health care workers who included 20 nurses,2 clinical officers, two gynecologists, six community health workers and three county directors of health. Purposive sampling was employed to select a diverse range of participants who could offer different perspectives and experiences and who were comfortable providing the information needed for the study.

Eligibility criteria were that the couples selected were of reproductive age and agreed to participate in the study, health care workers had been trained on cervical cancer screening and treatment and had worked in the clinics for at least six months, community health workers were stationed within local community, gynecologists and county health directors were deployed in the local counties. The researchers and the assistants took advantages of the health talks given in the mornings at MCH clinics to introduce themselves and the study then selected the participating couples The selection was done using simple systematic random sampling where the fifth couple was selected based on their sitting arrangement. Couples of reproductive age Seven in Murang’a, seven in Nyeri and six in Kerugoya were selected from MCH clinics. All the other participants were recruited based on their availability provided they met the criteria. The selected participants were further given explanation about the purpose of the study and confidentiality was maintained. After the researcher explained the purpose of the study and all their questions were answered, the information sheet and consent form were given for signing. The recruitment of the various study participants and data collection process took nine weeks in total (From 8^th^ April 2024 to 7^th^ June 2024)

### Data collection

Semi-structured interview guide questions were developed based on a literature review to address the objectives of the study and collect qualitative data. A pilot test was conducted in a neighboring county(Nyahururu) MCH clinic to test data collection instruments. Consent forms were given to the study participants which they all returned after signing to confirm their participation. The pre-testing of the data collection instruments was used to test the appropriateness of the interview guide questions and to provide the researcher with some early suggestions on the viability of the research. It also facilitated the researchers establish ways of building rapport with participants. The questions in both guides were open ended and probing, such as ‘Please explain how male partner can be involved in cervical cancer prevention and control?’, ‘Explain your role in cervical cancer prevention and control’, ‘What are the constraints/challenges encountered for males to support their partners during cervical cancer prevention and control?’

### Interviews

The semi-structured interview guide questions were posed to the participants one by one, followed by probing questions for more in-depth information. The interviews were primarily conducted in Swahili for community health workers, which all participants understood, while English was used for health professionals and directors. Each interview took place at a time convenient for the participant and lasted between 30 to 45 minutes. Field notes were taken, and audio recordings were made. The interviews were continued until data saturation was achieved, this was noted when all questions were answered, answers paraphrased to ensure clarity and unambiguity.

### Focus group discussions

Three FGDs were conducted involving couples from the selected three counties. The total number of participants were 40 (20 males and 20 females). The date was scheduled 7 days in advance and were notified to all the participants two times in between. Before the beginning of each FGDs session, the purpose of the study was explained to each couple participants. When all the participants agreed to take part in the discussion, the session was formally started. Considering adequate privacy, silence and adequate lighting, the site for FGDs was selected (one of the MCH rooms as there were no activities going on in the late afternoons). The participants were positioned in an arch shape inside the room. The principal investigator, as a moderator, and research assistant stayed at the front so that everyone would be visible to everybody in a comfortable manner. The main language for communication was Swahili. The participants were encouraged to interact with each other and some probing questions were used by the PI to make the discussion more effective. Each session lasted for about 45–75 minutes. All discussions were audio recorded. However, to supplement the transcripts, the research assistant also took notes. All the selected participants agreed to be audio recorded. The discussion was guided by laid down guide and continued until data saturation was achieved. This was noted when there was repetition of answers to various questions asked. The group members were assured of anonymity and confidentiality.

### Data analysis

Data was analyzed inductively. The analytical process included the following steps: verbatim transcription of recordings and field notes into Microsoft Word; confirmation and validation of transcripts; development of a thematic framework; coding of recorded conversations according to this framework; and charting and interpretation of the data (Vaismoradi et al., 2016). Microsoft Word was used for transcription, and emerging themes were recorded throughout this process. After transcription, the data was reviewed multiple times to internalize and refine key ideas. Data transcription and analysis were conducted after each interview, which helped to identify any missing information and prepare for the next interview. The data that was in Swahili from community health workers and FDGs was translated to English by the principle investigator as he understood both languages. The transcribed data were carefully reviewed for understanding the content and meaning. The researcher and his assistants went through the responses given by the participants and they confirmed them to be a true reflection of what they said. Codes were generated manually as per the second step of the analysis. Themes were developed by categorizing similar codes from the data and by creating specific groups. The themes that emerged were compared, briefly discussed and defined accordingly. Finally, a report was written to tell the story of the data, with direct quotes from some of the participants to support understanding of specific points thematically.

When coding, data was continuously reviewed, emerging patterns were noted and relationship between constructs was highlighted. Especially in FGDs, similar and contrasting viewpoints surrounding each theme was systematically analyzed by the authors.

### Trustworthiness of the study

The trustworthiness of the study was ensured by establishing confirmability, credibility, transferability, dependability and authenticity as illustrated in table 1.

**Table 1.** Trustworthiness strategies for qualitative data.

|  |  |
| --- | --- |
| Confirmability | • Member checking and peer debriefing to ensure that data accurately reflected the participant experiences. |
| Credibility | • A thorough collection of data ensured through conducting in-depth interviews.<br>• Member checking employed as a validation method.<br>• Direct quotations from the participants was incorporated.<br>• Field notes were diligently recorded during the interviews. |
| Transferability | • A detailed presentation of the study results was assembled.<br>• Purposive sampling was mainly employed, recognizing the limited scope for generalization in qualitative research. |
| Dependability: | • Peer debriefing sessions was held with proficient and experienced research assistants.<br>• A commitment to consistency was upheld throughout the data |
|  | collection process.<br>• Audit trail of data analysis procedures and recorded data. |
| Authenticity | • Data was audio recorded<br>• Use of direct quotes from participants to support themes and subthemes. |

### Ethical considerations

The initial step involved obtaining research approval from the University of KwaZulu-Natal’s Biomedical Research Ethics Committee (BREC), reference number BREC/00006580/2023. Additional ethical clearance from the country in which the study is based was secured from Mt. Kenya University’s Ethics Review Committee, reference number MKU/ISERC/3433. A license to conduct the research was granted by the National Commission for Science, Technology, and Innovation (NACOSTI) in Kenya, reference number 495117. Authority to collect data was also obtained from the relevant County Government Health offices in the study areas. Data collection commenced after securing written and signed informed consent from the participants, who had the right to withdraw at any time. Privacy and confidentiality were upheld, ensuring that data could not be traced back to individual participants. In all the stages of the study the ethical principles were upheld that included respect for persons, beneficence, integrity, objectivity and justice

## Results

### Characteristics of the participants

The total number of study participants were 73 comprising of 20 couples (20 males and 20 females) who were men and women of reproductive age,20 Nurses and two clinical officers who had been trained in cervical cancer screening and had worked in the clinics for at least six months, six community health workers, two gynecologists and three county directors of health who represented policy makers. The selected 20 couples participated in the focused group discussion while the rest participated in interviews.The mean age of the couples was 36.9 years for males and 30.5 years for females. Majority (70% n=28) of the couples had secondary education. The mean year for professional practice for nurses and clinical officers was 13 years. The participants were labelled as N_n,_ CO_n_, CHW_n_, G_n_, CDH_n_, FGD1-3_yn or xn_,where N Stands for Nurse, CO for Clinical officer, CHW for Community Health Worker G for Gynecologist, CDH for County director of Health, FGD1-3 for Focus Group Discussion one to three, n for serial number x for female member while y is for male member.

### Themes and subthemes that emerged from the data

Five key themes and eleven subthemes were generated from the data. The findings indicated that the male partner has several roles to play in transmission, prevention and control of cervical cancer. Table 2 Summarizes all themes and subthemes that emerged from data analysis. Findings are illustrated with quotations stemming from exact participant words from the original text

**Table 2.** Thematic analysis.

| **Objective** :To explore ways in which male partners are involved in cervical cancer prevention and control as perceived by couples and health workers. |  |  |
| --- | --- | --- |
| Question | Theme | Sub theme |
| What role do males play in their partners' cervical cancer prevention and control procedures? | 1.Financial and logistical support | 1.1Provision of financial support |
|  |  | 1.2Provision of logistical support |
|  | 2.Moral support | 2.1Emotional/psychological support |
|  |  | 2.2Positive reinforcement and open communication |
|  |  | 2.3 Advocacy |
|  | 3.HPV Transmission, Prevention and vaccination. | 3.1 HPV transmission and prevention |
|  |  | 3.2 HPV Vaccination |
|  | 4.Traditional and Cultural practices | 4.1Gender roles and norms |
|  |  | 4.2 Beliefs and practices |
|  | 5.Health education and health care provision(male as a health care provider) | 5.1Health education and awareness creation |
|  |  | 5.2 Health care provision |

### Theme 1: Financial and Logistical support

Male partners play an important role in encouraging their partners to utilize the availed cervical cancer prevention and control services. The support can be in form of material or financial support and sometimes logistical like incases of transport to where the health services are offered.

*Subtheme 1.1 Provision of financial support*

Male partner assisting with the costs of regular Pap smears, VIA or VILI tests, HPV testing and vaccinations, and any potential treatments can reduce financial burdens on the female partner.

The male partner can also ensure his insurance cover has provision for cervical cancer screening and treatment. When funds are not availed the woman may not go for checkups or may postpone the screening day until she gets the money. This was revealed by some of the participants; *“Majority of women are complaining of lack of funds even for registration”* (N_1_)

*“What about cervical cancer screening services? Is it free in all public hospitals in Kenya? “*(FGD1_y1_) The question was answered in affirmative by the researcher who also explained where to get the screening services.

When asked if male partners men supported their partners during cervical cancer prevention and control processes, they reported that some do financially and even accompany them to the clinic but majority of men prioritized their jobs more than their women’s health. They also reported that men feared going to hospitals and screening outcomes. Majority of men where community health workers were offering services lacked knowledge about cervical cancer therefore could not prioritize screening with their limited financial resources.

*“Most men do not prioritize cervical cancer since it is not them who are suffering, the pain is in the woman”* (CHW_1_)

*“Children tend to support their mothers morally and financially more towards seeking health services than their fathers”* (*CHW*_2_.)

*Subtheme 1.2 Provision of logistical support*

Male partner can organize for transport services to the health facility where screening and treatment of cervical cancer is done. He can also ensure that the domestic chores are taken care of when his partner go for screening and treatments of cervical cancer. Some of the participant’s responses pointed towards this subtheme;

*“Majority of male partners are disinterest on their women reproductive health, consider cervical cancer screening as women affair and business*” (CO_1_)

*“Some of my friends can’t attend these clinics you are talking about as they are overwhelmed by household chores including childcare and farm work”* (FGD2_x1_)

This indicated that if men could get interest in their partner cervical cancer screening processes they could relieve them some of the duties/chores.

### Theme 2: Moral support

By providing moral support, a male partner will create a safe and supportive environment for his partner, this will in turn help her to overcome the challenges of cervical cancer screening and treatment with more resilience. The moral support may entail emotional support, empathy, patience, positive reinforcement, advocacy and open communication.

*Subtheme 2.1 Emotional Support:* Offering emotional support, understanding, and empathy during the processes of cervical cancer prevention and control can reduce stress and anxiety for a female partner. Findings from the participant showed that male partners can support their female partners by accompanying the partner to the clinic, being concerned about the partner’s reproductive health e.g. by reminding her clinic days, Abstaining from sex for 6 weeks after cryotherapy treatment or thermal abrasion for a few weeks.

*“They are our husbands and they should be there when we are being checked the way they would like us to be there for them in case they are the one being checked”* (FGD2_x2_)

*“He should take me there for support incase am told I have cancer”* (FGD3_x3)_

*“Some of the husbands will come to the clinic to confirm when their wives tell them they have been instructed to abstain from sex for six weeks after cryotherapy”* (N_2_)

*Subtheme 2.2 Positive reinforcement and open communication:* Male partner support of healthy behaviors, such as regular screening and following medical advice, can boost a female partner’s confidence and motivation. Also Maintaining open and honest communication about fears, concerns, and feelings can foster a stronger bond and help both partners cope better. The respondents indicated that one way of positive support and reinforcement is avoiding judgment and stigma related to cervical cancer screening and being compassionate during the process of cervical screening and treatment.

“There *is lack of effective communication between women and their spouses as majority do not even inform their partners when coming to the clinic”* (N_12_)

*“Men will only come to hospital when they are severely sick, meaning screening services are alien to them”* (N_14_)

*“Some of the women tell me their spouses can’t accompany them for they fear the screening procedures or being informed that the results were positive for cervical lesions”* (G_1_)

On further probe about the fear, the gynecologist reported that some of the male partners are not comfortable watching their women being examined and also feared negative results. They are better off not knowing.

*Subtheme 2.3 Advocacy:* Male partner being a strong advocate for his woman healthcare needs, both within the relationship and with healthcare provision, can empower her and ensure she receives the best possible health services related to cervical cancer prevention and control.

Participants felt that male partner’s involvement and participation in cervical cancer prevention and control can be sustained through; -Screening being time sensitive i.e. not to take too long, teaching men the importance of their support, treating men well at home and inform them in advance, attaching a monitory value for attendance and having different set up for men in the clinical area.

*“Some of the males who come usually complain of slow moving ques and nurses going for tea breaks when they haven’t been served”* (N_7_)

*“There was a time I accompanied my wife for our child immunization and we were clumped together with women who were breastfeeding others very pregnant and the space was small, I felt so uncomfortable’*(FGD2_y2_)

The support that should be provided to couples and community to ensure sustained male partner involvement in cervical cancer prevention and control include Communication on the health issues including those affecting men, Empowering women to be less dependent on men financially. Government and NGO’s to fund screening drives, ensure accessibility to cervical cancer screening facilities, educate community Health workers, Pap smear and other cervical cancer diagnostic procedures to be free or affordable. More information of HPV vaccine safety should also be disseminated. This was reported by health care providers. One of the county director of health reported that laid down guidelines are essential to assist male partner involvement in cervical cancer control and treatment.

*“With proper guidelines I think ways can be organized to involve the male partner in cervical cancer screening even if it involves integrating it with prostate cancer screening and educating them about cervical cancer”* (CDH_2_).

### Theme 3: HPV Transmission, prevention and vaccination

Ninety-seven percent of all cervical cancer disease occurrence are related to HPV.This virus is mainly sexually transmitted meaning male sexual partner are the main vehicle for its transmission. Male parents also have a role to play in ensuring their children are vaccinated against HPV before their first sexual debut.

*Subtheme 3.1 HPV transmission and prevention*

Men are usually asymptomatic carriers of HPV, meaning they may not exhibit any symptoms but can still transmit the virus to their female partners. HPV can be transmitted through skin-to-skin contact during sexual activity, even when _there_ are no visible genital infections. Being faithful to one non infected partner and using condoms consistently during sexual activity can significantly reduce the risk of HPV transmission. Participants revealed some of the ways male partners can be involved in HPV Transmission and prevention to include avoidance of multiple sexual partners and wearing condoms persistently for those with multiple sexual partners. Some of the group members were not aware that cervical cancer can be transmitted sexually. *“Cancer is not an infection how can it be transmitted sexually”.* (FGD3_y4_)

*“Do you mean those women with cervical cancer got it from their partners?”* (FGD2_x5_)

The investigator explained how HPV is transmitted sexually and its association with Cervical cancer.

Health care workers also reported lack of knowledge among their clients regarding transmission of cervical cancer.

*“Many clients coming to our clinics have no basic facts regarding cervical cancer and its prevention”* (N_8_)

*“Majority of men are not even aware that Cervical cancer can be sexually transmitted”* (N_10_) *“Majority of male partners are disinterest on their women reproductive health, consider cervical cancer screening as women affair and business*” (CO_1_)

*Subtheme 3.2 HPV Vaccination*

HPV is primarily a sexually transmitted infection, and the vaccine is most effective when administered before sexual activity begins (Markowitz and Schiller, 2021). Parents including fathers are responsible in ensuring their children are vaccinated against HPV on time. Some of the roles the respondents revealed that male partner can promote HPV Vaccination included, allowing the children below 14 years to be vaccinated against HPV, encouraging other men to take their children for HPV vaccination, taking his children for HPV vaccination, dissuading myths and misconception regarding HPV vaccination, vaccinating his children against HPV, taking his partner for HPV testing and encouraging his partner to go for HPV testing.

There were a lot of myths, misconceptions and lack of awareness regarding cervical cancer, screening and HPV vaccination among the focused discussion group members as the following reported verbatim illustrate;-

*“Some people don’t have girls and they are advocating for this vaccine on high note, how stupid are we to forget that the other vaccine for polio 4 years ago? Were it not for the catholic church we could be singing another song now, to hell with their vaccines, mine NO! “*(FGD2_y5_)

*“What should be done if you didn’t get the vaccine during childhood or ages mentioned? “*(FGD1_x5_)

*“In which facility within the locality is the vaccine available? “*(FGD3_y6_)

These questions and concerns were answered comprehensively by the researcher and the assistants.

Some of the participants had positive attitude towards the vaccine *“Mine is vaccinated…two doses, this vaccine is safe…cancer is painful and expensive to treat, kindly let your daughter be vaccinated and she will be safe as mine. “*(FGD1_x5_)

*“Mine is vaccinated awaiting the second dose “*((FGD3_y5_)

### Theme 4 Traditional and Cultural practices

Cultural and Traditional practices may significantly influence male partner involvement in cervical cancer prevention. This may include Gender Roles and Norms and patriarchal nature of many societies.

*Subtheme 4.1 Gender norms and roles:* In societies with patriarchal structures, men may hold dominant positions, leading to a lack of involvement in women’s health issues. Cultural norms may dictate that men have the primary decision-making power regarding healthcare, limiting women’s autonomy. Participants reported that men should lead by example and stop associating accompanying their wives to reproductive health clinics with weakness.

*” It is difficult for men in this community to accompany their wives here due customs and traditional perceptions*” (N_17_)

“*Most men are busy looking for money and drinking so they are not readily available*” (N_3_) *“Some of the males who come usually complain of slow moving ques and nurses going for tea breaks when they haven’t been served”* (N_7_)

*Subtheme 4.2 Beliefs and practices:* Reliance on traditional healers for health concerns can delay seeking professional medical care. Stigma and Shame associated with sexually transmitted infections including cervical cancer can prevent men from discussing sexual health concerns with their partners or seeking medical advice. Some of the practices that the participants identify that can undermine cervical cancer prevention and control included polygamous relationships and wife inheritance.

*“Some male partners in this community don’t believe in conventional medicine and relies on traditional herbs meaning they can’t take their spouses for cervical cancer prevention and control health services”* (CHW_3_)

“*Some males will be embarrassed to accompany the wives to maternal and child health clinics”* (N_5_).

*“Some men will never appear in these clinics based on the sitting arrangement in the waiting area*” (N_11_)

### Theme 5 Health education and health care provision

Male partners can play a significant role in promoting health education and facilitating healthcare provision for cervical cancer prevention and control. By actively being involved in health education and healthcare provision, males can contribute to a healthier and more informed society, reducing the burden of cervical cancer within the communities they live.

*Subtheme 5.1 Health education and awareness creation:* this involves being in the front line in educating the family and community about cervical cancer, Community mobilization in cervical cancer prevention and control processes and learning and teaching on how to prevent and control cervical cancer. It may also involve provision of financial and other resources anytime his partner is attending sessions that are educating about cervical cancer.

“we *can educate the community on the importance of male participation in cervical cancer prevention through Educational campaigns involving the media*” (CO_2_)

*“It will be more impactful if males are informed by their peers or friends on the importance of cervical cancer screening and the need to support them holistically”* (G_2_)

“*In my opinion sex education from junior schools to tertiary educational centers will ensure men are properly informed about cervical cancer and how to mitigate against it”* (N_6_)

*Subtheme 5.2 Healthcare provision:* Participants reported that this can be through ensuring males are among the trained health services providers in relation to cervical cancer, Screening women for cervical cancer, treating women with cervical lesions appropriately and referring women to higher/advanced care in case they can’t be treated/managed locally. Policy makers reported that sometimes it’s challenging to ensure there are male health care workers especially nurses to offer cervical cancer screening and treatment services.

*“It is not possible to focus on male trainees as the we train those who are already in the clinics and they are mainly women by default”* (CDH_1_). *“We have very few male nurses in our MCH as majority are in the wards. May be it may help to have more male nurses in those clinics to encourage men to bring their wives and children”* (CDH_2_).

*“Our healthcare professionals currently lack adequate skills to effectively handle male clients. I recommend that we incorporate both theoretical and practical training simultaneously to enhance their competency in managing male clients. This approach will better equip them to encourage men to support their wives in reproductive health matters.”* (CDH_3_).

## Discussion

Cervical cancer should be viewed not just as a health issue affecting individual women but as a broader social issue that requires greater public awareness of men’s roles in both transmission and prevention (Moodley and de Vries, 2016). Engaging men through advocacy, education, and participation in cervical cancer screening, while respecting socio-cultural norms, can help them make informed decisions about their partners’ cervical cancer screening. In many African contexts, men often hold dominant positions within the family (Adegboyega A et al., 2019). This may influence the decision making processes that may affect uptake of reproductive health services including cervical cancer prevention and control.

The study focuses on the opinions of couples, health professionals, community health workers and health policy makers on male roles in cervical cancer transmission, prevention and control.

Some of the roles and responsibilities for male partners in transmission, prevention and control of cervical cancer identified in this study included-;

### Financial and Logistic support

The study found out that male partners are involved or need to participate in financial and logistic support during their partners’ processes of cervical cancer prevention and control health services. This involves provision of money for fare to the clinic, money to pay for registration in the clinic

Providing financial support for services, covering the cost of household help when a partner visits the clinic, and purchasing medications and other supplies needed after screening are crucial. Previous research has highlighted that in Uganda, gender power dynamics are patriarchal, with men traditionally controlling family finances and access to healthcare (Mwaka et al., 2013; Mutyaba et al., 2007). Although the available data does not allow for strong conclusions about the impact of men on cervical cancer screening uptake, involving men in the screening process could be beneficial. Their involvement might help facilitate women’s attendance through emotional and financial support and ensure proper follow-up (Mwaka et al., 2013 In his dissertation on factors influencing uptake of VIA/VILLI cervical cancer screening and treatment in Kenya Mwangi reported that male partners were a factor in determining the uptake of the services as he was the one to provide the funds for transport and the hospital fee(Mwangi, 2015)

### Moral support

According to the study findings, supporting a partner through cervical cancer screening involves accompanying them to the clinic, showing concern for their reproductive health by reminding them of clinic days, abstaining from sex for six weeks after cryotherapy/thermal ablation treatment, and avoiding being judgmental to prevent stigma. It also includes being compassionate during the screening process.

In a study conducted in Sub-Saharan Africa, women indicated that a lack of spousal support was not a significant barrier to accessing screening services. However, some women expressed concerns that their male partners might abandon them or refuse to cover the costs if they tested positive for cervical cancer or HIV, suggesting that gender power dynamics play a role (Lim and Ojo, 2017). This finding is supported by a randomized controlled trial in Uganda, which found that women referred for colposcopy after a positive screening test were more likely to return for the procedure if their partners were involved in the process (Mutyaba et al., 2007).

### HPV transmission and prevention

A significant association exists between cervical cancer and repeated human papillomavirus (HPV) infections over time (Dillner and Brown, 2004). HPV can be transmitted from an infected man to an uninfected woman through sexual contact (Denny et al., 2010). Once oncogenic HPV is transmitted, cervical cancer develops gradually, with symptoms potentially taking many years to manifest (Dillner & Brown, 2004). This indicates that while women bear the primary burden of the disease, male sexual partners contribute to the transmission of the virus to their female partners (Moodley and de Vries, 2016).

A study conducted in India on the impact of male behavior on cervical carcinogenesis found that husbands’ premarital and extramarital sexual relationships increased the risk of their spouses developing cervical cancer by a factor of 6.9. Additionally, sexual intercourse with uncircumcised men or those circumcised after the age of one year was associated with a 4.1-fold increased risk of cervical cancer (Agarwal et al., 1993).

Although young boys should be vaccinated against the HPV, the HPV vaccine has often been labeled the “girl vaccine” (Mishra and Graham, 2012). Even within the dissemination of health and medical information, the male role in cervical cancer prevention is not clearly emphasized. For instance, a 2015 editorial in the South African Medical Journal argued for vaccinating girls without mentioning the role of males (Botha et al., 2015; Moodley and de Vries, 2016). From an objective health perspective, this omission effectively normalizes the lack of male involvement. Additionally, the vaccine is often marketed as a cancer prevention measure, avoiding discussions about its connection to sexually transmitted infections, which could implicate male partners (Mishra & Graham, 2012; Davies and Burns, 2014). Media representation has also been gendered, focusing predominantly on girls and women while leaving boys and men largely out of the conversation (Giuliano et al., 2007; Thompson et al., 2020). However, there is a promising shift: a recent study on social marketing strategies found that preteen boys in an intervention region were 34% more likely to get vaccinated compared to those in regions without such interventions (Cates et al., 2014).

When the role of men in the heterosexual transmission of HPV is under-recognized and HPV is seen exclusively as a female issue, the medical system—including initial vaccine trials and marketing focused primarily on girls—has effectively delayed or denied male accountability (Moodley and de Vries, 2016). This is further supported by a study in the U.S. where parents expressed willingness to vaccinate their adolescent sons, but had not done so because healthcare providers had not recommended it (Donahue et al., 2014). This suggests that medical practitioners may be avoiding discussions about vaccinating boys, thereby reinforcing the misconception that HPV is solely a female concern. Similarly, Canadian nurses have celebrated the HPV vaccine within the context of cervical cancer and traditional feminine roles (Mishra & Graham, 2012). These reports highlight the significant feminization of HPV vaccination and testing, underscoring the need for change to recognize the role of males in HPV transmission and prevention.

From this study findings majority of male partners were not aware of the relationship between cervical cancer and HPV infection meaning that they couldn’t associate their sexual behavior with cervical cancer transmission. This is supported by the study in Midwest Uganda where men were willing to support cervical cancer screening and treatment for their wives and daughters vaccination after being informed about cervical cancer (de Fouw et al., 2023).

There was a lot of myths and misconception amongst focused group discussion members regarding cervical cancer and HPV vaccine, but it was encouraging to note that those who had the knowledge about the vaccine were taking or willing to take their children for HPV vaccination. This was consistent with a study done by Robert A Bednaczyk where he addressed five key myths-Pap smears are sufficient to prevent cervical cancer; HPV vaccination is not safe; 11–12 years of age is too young to vaccinate, HPV vaccination is not needed since most infections are naturally cleared by the immune system (Bednarczyk, 2019). From the study male’s roles in prevention of cervical cancer through HPV prevention entails; Allowing the children below 14 years to be vaccinated against HPV, taking his children for HPV vaccination, encouraging and facilitating other men to take their children for HPV vaccination, dissuading myths and misconception regarding HPV vaccination, Avoiding Multiple sexual partner and encouraging and taking their partner for HPV testing.

### Traditional and Cultural practices

Traditional and cultural practices can potentially influence male partner involvement in cervical cancer prevention and control processes. These practices can create barriers to understanding, participation, and support for women’s health issues including reproductive health. Low knowledge, socio-cultural practices and inadequate and inappropriate services for men negatively affects utilization of health services and impairs support for their wives’ service uptake(Kura et al., 2013).The study identified male masculinity as a barrier in male partner support towards cervical cancer prevention in that some male partners associated accompanying their women to MCH clinics as a weakness. Some men were also reported to have low opinions on conventional health services and were relying more on traditional herbs and other remedies. The male masculinity where accompanying women to the hospital is perceived as a weakness should be discredited. This can be through campaigns and mobilizing the gatekeepers in the community to advocate for male support during cervical cancer prevention and control strategies. The findings are supported by a study in Uganda where cervical cancer screening among married women was significantly associated with intimate-partner factors; women’s education attainment, intimate-partner’s emotional and financial support. The intimate-partner’s factors associated with cervical cancer screening point to traditional marital roles of male partner dominance. Efforts to encourage men’s participation through community education was recommended. (Isabirye, 2022)

### Health education and awareness creation

The findings showed that male partners should be in the front line in educating the family and community about cervical cancer. He should also mobilize the community resources to educate members about cervical cancer. They should also learn and teach family members on how to prevent and control cervical cancer. Additionally, partners can offer financial and other resources whenever their partner is attending educational sessions on cervical cancer. This approach aligns with findings from Rural Ghana on male support for cervical cancer screening and treatment where males were willing to support their partners socially, financially, materially and emotionally but lacked education about cervical cancer. (Binka et al., 2019)

In another study on Uyghur women in Xinjiang, China, findings suggested that lack of knowledge about cervical cancer significantly contributed to its high incidence in the population. The study concluded that comprehensive health education to raise awareness about cervical cancer and HPV is crucial for reducing incidence and mortality rates among Uyghur women, potentially even more so than prophylactic vaccination (Abudukadeer et al., 2015). This can be enhanced by creating awareness within the community about cervical cancer and how it can be prevented and controlled.

### Health care provision

Male partners are integral part of the health care system, serving as trained professionals involved in cervical cancer prevention and control. They are responsible for screening, treating, and referring women to her facilities appropriately. To increase the number of male health service providers offering cervical cancer screening, additional on-the-job training is essential. This recommendation aligns with findings from a study on Clinical Training of Visual Inspection with Acetic Acid (VIA) in Kenya, which advocated for expanding training for healthcare providers in VIA and cryotherapy (Maina, 2023). This expansion should include male health service providers working in maternal and child health clinics.

### Limitations of the study

Being a qualitative study and being carried out in only public health facilities in central Kenya the study may not be generalized to represent the general population. Further research need to be done using a different methodology and to include private and mission health facilities to compare the findings.

## Conclusion

Based on couples, health workers and policy makers perceptions on the roles of male partner in transmission and control of cervical cancer, male partners are not yet fully supporting their partners in preventing and controlling cervical cancer. This can be attributed to their lack of awareness about the disease, financial constraints, cultural and social attributes and sometimes health care system factors. By addressing men’s knowledge gaps, promoting responsible sexual behaviors, encouraging open communication about cervical cancer through acceptable cultural/tradition set ups, improving the health care services, males will support their female partners in the journey of cervical cancer prevention and control.

Cervical cancer should not be viewed solely as a women’s health issue, but rather as a broader public health concern that requires a deeper understanding of the roles male partners play in its transmission, prevention, and control.

The key roles of male partners identified in the study include providing financial, logistical, and moral support, engaging in health education and awareness efforts about cervical cancer, challenging and adjusting harmful social norms and traditional beliefs, participating in healthcare provision (including as part of the medical staff), avoiding multiple sexual partners, and supporting HPV vaccination.

### Recommendations

Policymakers should integrate male partner involvement into existing national health programs and campaigns, ensuring that messages and strategies are effectively tailored to reach men. They should also promote awareness and education among men about cervical cancer and emphasize the importance of their role in prevention, including supporting HPV vaccination and encouraging regular cervical cancer screening for their partners.

Program implementers should develop culturally sensitive and engaging interventions that address the specific needs and concerns of men within the target population. By utilizing a variety of communication channels, they can effectively reach men and empower them to take an active role in cervical cancer prevention and control. This includes providing men with the necessary information, skills, and resources to support these efforts.

## Acknowledgements

The author would like to acknowledge Prof. N.G. Mtshali and Dr.P. Mbeje for their full support as supervisors in writing this manuscript. Acknowledgement is extended to all the study participants for their cooperation and full engagement.

## Data availability

Data sharing is not applicable to this article as no new data were created or analyzed in this study.

## Disclaimer

The views and opinions expressed in this article are those of the author and do not necessarily reflect the official policy or position of any affiliated agency of the author.

